# Validation of AI-based software for objectification of conjunctival provocation test results in routine examinations and clinical studies

**DOI:** 10.1101/2022.03.23.22272791

**Authors:** Yury Yarin, Alexandra Kalaitzidi, Kira Bodrova, Ralf Mösges, Yannis Kalaidzidis

**Author notes:** Correspondence to Yury Yarin and Yannis Kalaidzidis.

## Abstract

**Background:** Provocation tests are widely used in allergology to objectively reveal patients’ sensitivity to specific allergens. The objective quantification of an allergic reaction is a crucial characteristic of these tests. Due to the absence of objective quantitative measurements the conjunctival provocation test (CPT) is a less frequently used method despite its sensitivity and simplicity. We developed a new method AllergoEye based on AI for quantitative evaluation of conjunctival allergic reactions and validated it in a clinical study.

**Methods:** AllergoEye was implemented as a two component system. The first component is based on an Android smartphone camera for screening and imaging the patient’s eye and the second one is PC based for image analysis and quantification. For the validation of AllergoEye an open-label, prospective, monocentric study was carried out on 41 patients. Standardized CPT was performed with sequential titration of grass allergens in 4 dilutions with the reaction evaluated by subjective/qualitative symptom scores and by quantitative AllergoEye scores.

**Results:** AllergoEye demonstrated high sensitivity (98%) and specificity (90%) as compared with human-estimated allergic reaction. Tuning cut-off thresholds allowed to increase the specificity of AllergoEye to 97%, where the correlation between detected sensitivity to allergen and sIgE CAP-class becomes obvious. Strikingly, such correlation was not found with sensitivity to allergen detected by subjective and qualitative symptom scores.

**Conclusion:** The clinical validation demonstrated that AllergoEye is a sensitive and efficient instrument for objective measurement of allergic reactions in CPT for clinical studies as well as for routine therapy control.

## 1. Introduction

Provocation tests in allergology play an especially important role in the clinical cases of patients with allergic symptoms who have negative skin reactions and negative Carrier-Polymer-System (CAP) test results. The positive provocation reactions in this case are an objective proof of allergy. On the other hand, negative results of provocation in patients with positive skin tests and/or CAP without active allergic symptoms can reveal a sensitization without clinical relevance. Therefore, with both positive and negative outcomes, provocation tests support the decision on the necessity of therapy.

Among other provocation tests the conjunctival provocation test (CPT) is simpler than others without compromising accuracy and sensitivity [1,2]. In the last years the CPT is used for diagnosis as well as for therapy control and is becoming a crucial instrument of clinical studies [3,4,5]. However, despite a hundred years of CPT history [6,3], its role is still limited due to the main drawback - the subjective character of the evaluation of outcomes. Nevertheless, the CPT has the same level of clinical importance as the nasal provocation test, which includes an objective instrumental evaluation of outcomes [1,3,7]. Multiple clinical studies demonstrated a high level of concordance between these two methods [8, 9, 10, 11]. Up to date the main method of evaluating the CPT is a summation symptom score (SSS) [2]. SSS is the sum of categorically (none (0), mild (1), moderate (2), severe (3), intolerable(4)) estimated symptoms, part of which are subjective patient estimation of allergy symptoms (like eye itching, irritation and tearing) and eye redness subjectively estimated by medical staff (observers) carrying out the test [3,11,6].

Modern evidence-based medicine requires quantitative reproducible evaluation of test results for diagnostics as well as for assessments of the effectiveness of therapy carried out. To overcome the CPT drawback, several approaches were proposed for the quantification of CPT-outcomes [4, 12, 13, 14]. The readout of quantitative CPT methods is based on eye redness measurement on digital images. The redness quantification can be divided into two approaches. The first approach is based on the calculation of the apparent area of vessels in the sclera [12, 15]. Therefore, vessels were 1) contrasted, 2) segmented and then the relative area of sclera covered by segmented vessels was taken as a redness score. However, the partial observation of the sclera and different eye ball orientation toward the camera on sequential observations could degrade the accuracy of the method. Additionally, diffuse redness, which is related to the widening of capillary that could not be resolved on non-microscopic images, is not captured by this method. The second approach calculates the redness for each pixel [4, 14]. The mean redness of the sclera [4] or histogram of redness distribution [14] was then used as a redness score. In this case, one has to note, that there are multiple definitions of redness [16, 4, 17] that could lead to different sensitivities of the method. The crucial problem of per-pixel redness estimation is its dependency on the reproducible color balance of the environment illumination and the automated white balance of digital images. The limited application of all mentioned methods is either due to semi-automated image analysis [4, 18, 12] or due to absence of automated correction for illumination color changes (white balance) [14] or both of them [4, 18, 12, 14].

At the end of the last decade, the breakthrough in the Deep Learning approach brought Artificial Intelligence methods into wide practice. In medicine it was applied in diagnostics, image reconstruction in radiology modalities, emergency care devices etc. [19,20,21]. However, its usage in allergology is still limited. Recently we developed a new approach for the qualitative evaluation of CPT results for diagnostics of allergic rhinoconjunctivitis and implemented it in high-throughput software platform AllergoEye, which comprises of Deep Neural Networks for automated image analysis and symptom evaluation [17]. For the implementation of AllergoEye in broad clinical practice a validation study for quantitative estimation of sensitivity and specificity of the method was required. In the presented work we demonstrate the results of the clinical validation and efficiency of AllergoEye on a cohort of patients with grass pollen allergy.

## 2. Methods

### 2.1 Approvals and ethics

The study was approved by the Ethics Committee (IEC) of the State Chamber of Physicians of Saxony, Germany (IEC number EK-BR-111/21-1). The study was conducted in accordance with local regulations, the International Conference on Harmonization of Technical Requirements for Registration of Pharmaceuticals for Human Use (International Conference on Harmonization (ICH)–Good Clinical Practices (GCP)) and the Declaration of Helsinki [22].

### 2.2. Study design

This open-label, prospective, monocentric study was carried out in the Clinic of Otorhinolaryngology and Allergology Dr. Yury Yarin (Dresden, Germany) in 2021. Female and male patients between 18 and 75 years of age were included in this study. The study on these grass pollen-allergic patients was performed outside of the grass pollen season and consisted of 2 visits. Inclusion in the study took place after the patient signed the informed consent prior to any study procedures being performed. For participation in the study the patients were required to have a history of moderate or severe seasonal allergic rhinoconjunctivitis (AR) with or without seasonal controlled asthma during the three grass pollen seasons in the past. For objectivities of the allergy as criteria for the inclusion in a study, a skin prick test and grass pollen–specific IgE (sIgE) antibodies were performed. A wheal diameter 2≥3 mm for a grass pollen allergen solution and sIgE antibodies > 0.01 kU/L were the main necessary condition for study inclusion. Exclusion criteria were adapted from: guidelines for daily practice [3] (see attachment 1).

During the first screening visit, the allergic anamneses was collected, the inclusion/exclusion criteria were checked. After that the skin prick test was performed and a blood sample for the antibody was taken. During the second visit, CPT was performed and estimated by summation symptoms scores (SSS) [18] which were calculated at the end of the test by medical personnel.

Simultaneously to the CPT procedure the results of patient’s reactions on each applied dilution were additionally recorded by AllergoEye. The results of the CPT and of AllergoEye were joined into a merged database and statistical analysis was performed.

### 2.3. Titrated CPT Procedure

The CPTs and AllergoEye recording were conducted by the same investigator. Standardized lyophilized allergen extracts for CPT with concentration in Histamine Equivalent Potency units (HEP) 30 HEP/ml containing grass group supplied by Laboratorios LETI S.L. Madrid, Spain were used. Reconstitution CPT stock solution was performed by drawing 4.8 mL from a vial with 5.0 ± 0.2 mL of solvent and dispensing into a vial with grass extract. Further dilution steps were 1:1000, 1:100, 1:10. A diluent will serve as negative control. The CPT was performed by the following protocol: At the first step of the CPT, a negative control solution (without allergens) is applied to the right eye. In case of a negative reaction after 5 minutes, an allergen solution is applied stepwise to the left conjunctival sac in predefined increased concentrations 1:1000, 1:100, 1:10 and stock solution. Between each application a 5 minutes interval was held. The symptoms were documented in a standardized form [18] (see Table 1 in Suppl.Fig.3) in 5 minutes after allergen application. The four symptoms of tearing, itching, irritations and redness were analyzed. Each of these symptoms was estimated and categorized on a scale from 0 to 3, whereby, 0 = no reaction, 1 = mild reaction, 2 = moderate reaction, and 3 = strong reaction. Total score SSS was summarized from the meaning of all 4 symptoms subjective (itching, irritation and tearing) and objective (redness evaluated by medical staff) and built the value ranging from 0 to 12. The protocol was stopped when either medical personnel detected redness of the treated eye or maximum allergen concentration was achieved. Finally the allergen-provoked eye was treated with antihistaminic eye drops (Ketotifen).

### 2.4. AllergoEye - measurements and protocol

During the CPT we described above, both patient’s eyes were imaged 5 min after each provocation, which was done on the left eye, with sequentially increased concentration of the grass CPT solution. Control images were taken also before the test and after the treatment of the right eye with a control solution (test solution without allergen). The eyes were imaged at 3 positions (looking straight forward, to the right and to the left) for maximum coverage of the sclera. To control the illumination condition, the images were taken with a special mask (see Figure 1A) with continuous white LED illumination. At the end of the test the images were sent to a PC where the redness was evaluated by AllergoEye software.

**Figure 1.**
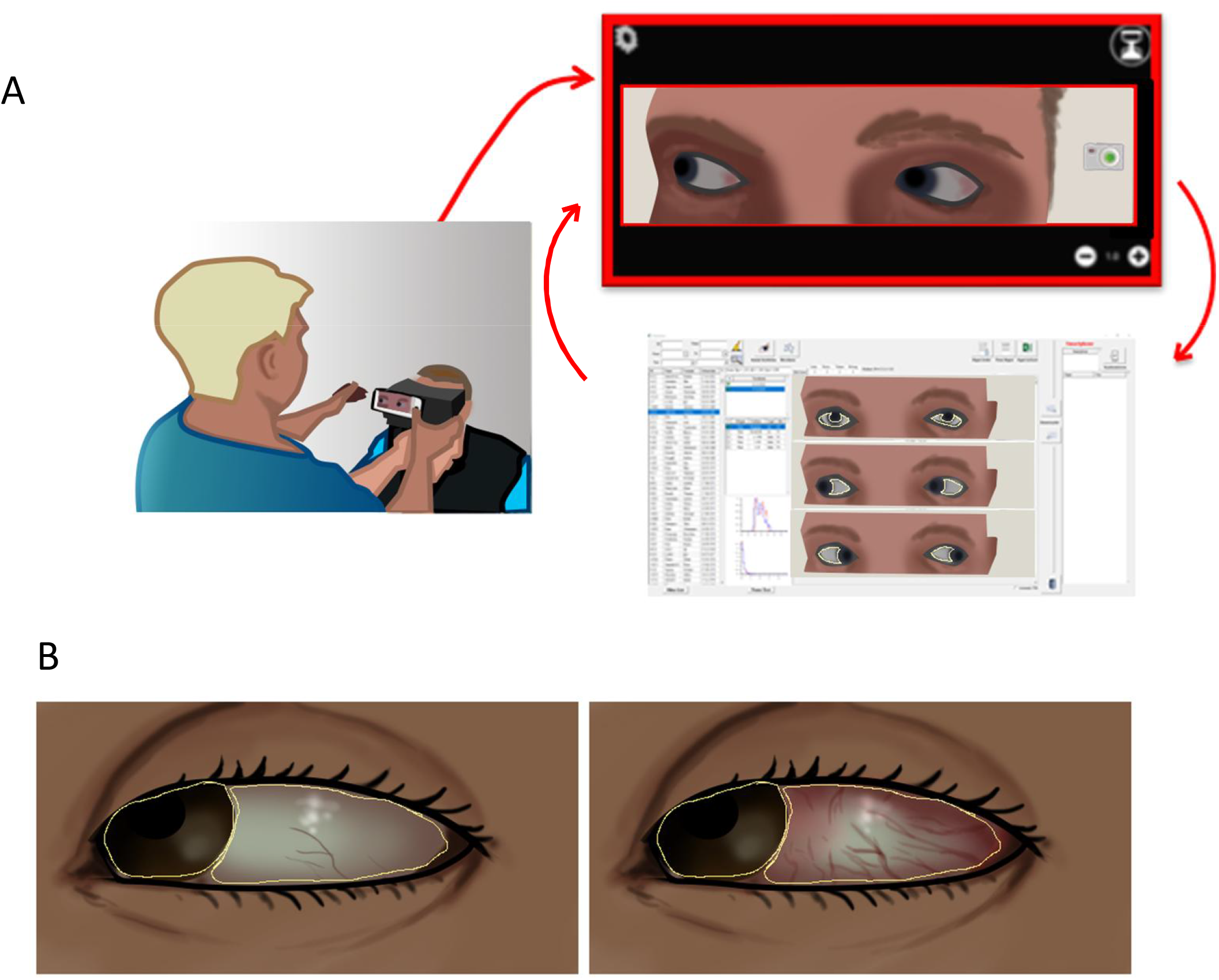
Workflow of PCT evaluation by AllergoEye. **A**. (left) Nurse makes image of patient’s eyes by smartphone using AllergoEye mobile app (right), then image (top) is wireless transferred to PC-based AllergoEye application (bottom right), the result of analysis is transferred to mobile application for control. **B**. Sclera and iris are segmented by neural network (contoured in yellow). Sclera redness before (left image) and after (right image) provocation is shown.

### 2.5. AllergoEye - technical description

AllergoEye was implemented as a two component system. One part is based on an Android smartphone HONOR 20 Pro (HUAWEI) and uses the build-in high-resolution photocamera for imaging the patient eye’s reaction to the allergen (Figure 1a). The second part is a PC based software system, including the patient database, the communication module for exchanging data with multiple smartphones and a deep neural network for image analysis and redness evaluation. In short, patient’s data and measurement details (allergen, dilution, exposure time) are transferred from the PC to the mobile phone. The nurse acquires images of two eyes and wirelessly transfers them to the PC. There, the deep neural network is used to recognize and segment the iris and sclera. Additional control of coupling of iris and sclera as well as the presence of two open eyes was performed to exclude segmentation artifacts. We proposed a new method of per-pixel redness of sclera (*R*) evaluation as:

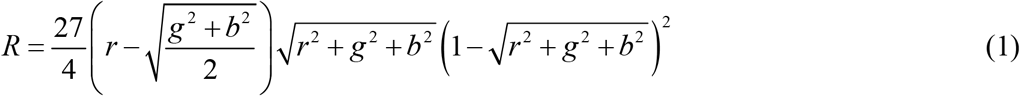

where *r, g,b* are normalized red, green and blue intensities respectively. The mean redness and distribution of redness in the sclera are calculated for both treated and non-treated eyes from all 3 eye positions. To take in account the inhomogeneity of the sclera reaction to the allergen (see Figure 1b), the distribution of squared gradients of redness with 4 spatial steps were calculated.

Despite of usage of a special mask (see Figure 1a), in some cases the changes of illumination in the room during the examination led to the shift of white color balance of images (compare right and left images on Figure 1b). To make our measurement robust to the illumination change, we used the sclera of the untreated (right) eye as an internal control. For this end, the redness distributions of both eyes were normalized (shifted) in such a way, that the redness distribution of the right eye on test and control images had a maximum possible overlap. After such normalization the difference between redness of the left eye in test and control conditions were used as AllergoEye scores.

### 2.6. Statistical Analysis

For significance estimation the Student-t test was used. * denotes p_value_ < 0.05, ** denotes p_value_ < 0.01, *** denotes p_value_ < 0.001. Statistical analysis and graph generation was performed by model analysis software FitModel (http://pluk.mpi-cbg.de/projects/fitmodel).

## 3. Results

First we characterized the distribution of patients in the study (N=41) by eye sensitivity to the grass allergen. As shown in Figure 2, most of the patients (n=16; 40%) exhibited a reaction at allergen dilution 1:10, whereas only 4 (10%) were sensitive to dilution 1:1000. It was found that 3 (8%) patients did not reveal any eye reaction for provocation at highest concentration (stock solution) despite anamneses record of moderate to severe rhinoconjunctivitis.

**Figure 2.**
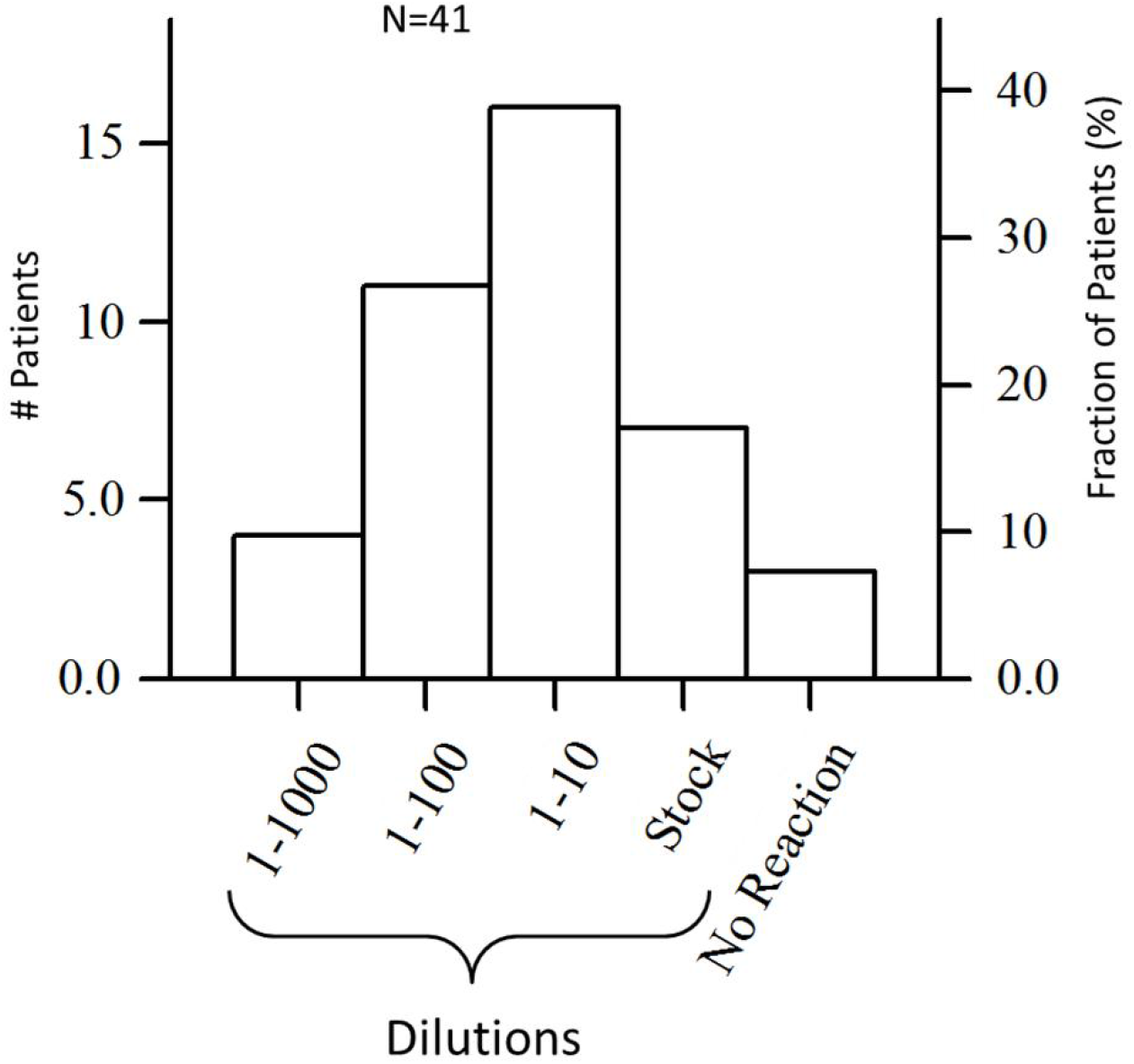
Distribution of 41 patients by sensitivity defined as allergen dilution, where allergic reaction was detected as either SSS 2 ≥ 3. Left axis is number of patients; right axis is of percent patients.

Next we characterized AllergoEye’s sensitivity/specificity. For this end we built a scatterplot for 114 measurements (Figure 3a, blue points). Each point represents one measurement, where the x-value is human-estimated redness score (from 0 to 3) and the y-value is an AllergoEye score (from 0 to 3.7). Then we gradually changed the threshold value from 0 to 3.7, and for each threshold value we labeled the measurements as positive (above threshold) or negative (below threshold). As the ground-truth labels we took human-assigned scores. Therefore, a positive value was labelled as true-positive if it corresponds to human-assigned non-zero score; otherwise it was labelled as false-positive. The true- and false-negative labels were assigned in a similar way. From these labels we constructed a ROC-curve for AllergoEye, where for each threshold value we calculated

**Figure 3.**
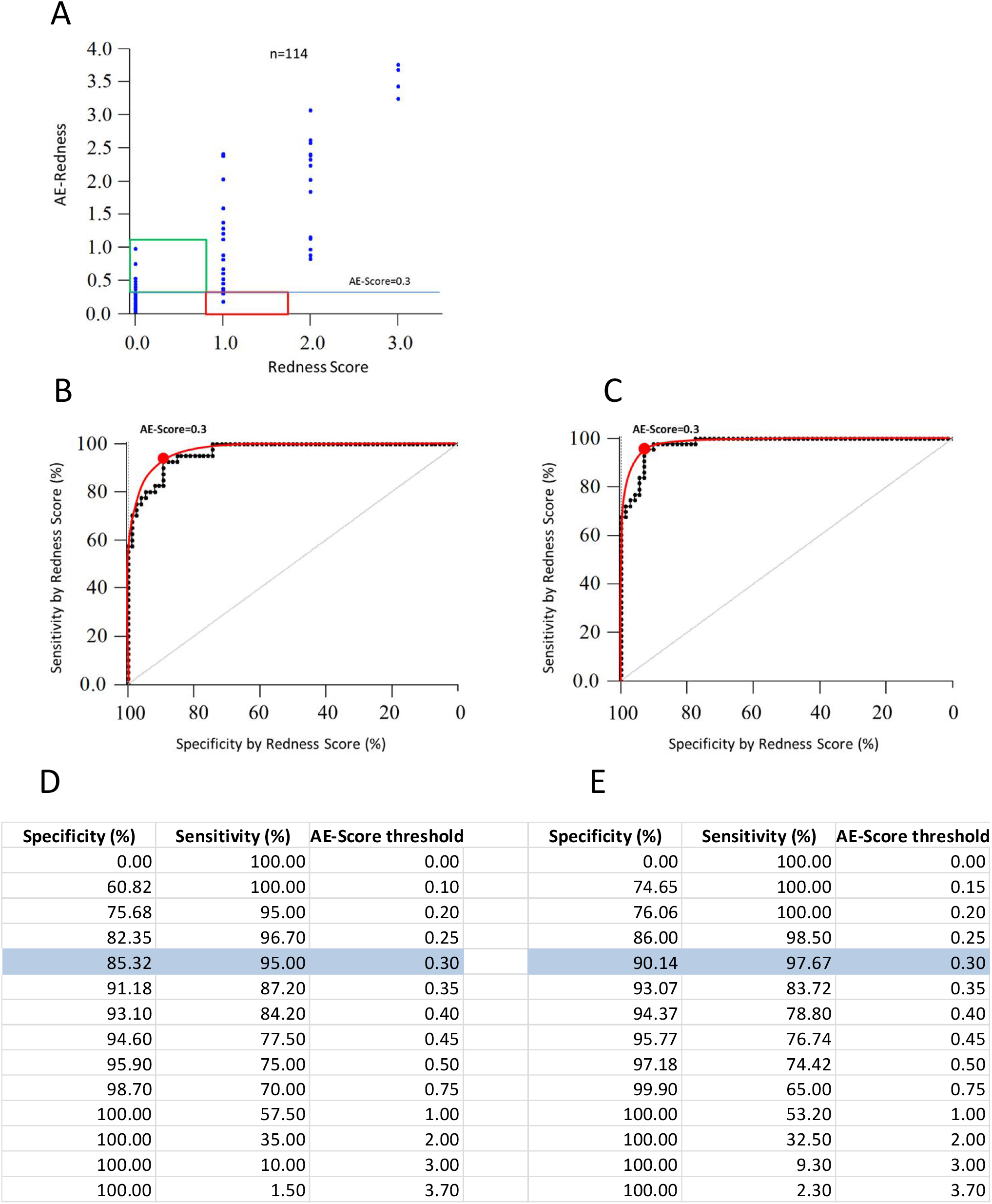
**A**. Scatter plot of 114 measurements (blue dots), which were performed on 41 patients to reveal dependency between redness score estimated by medical staff (horizontal axis) and redness evaluated by AllergoEye (vertical axis). Horizontal cyan line denotes threshold equal 0.3 (see text). Red and green rectangles highlight measurements that “false negative” and “false positive” respectively. **B, D**. ROC curve (**B**) (Sensitivity vs. Specificity) as defined by threshold value gradually changing from 0.0 to 3.7 and its table presentation (**D**). Experimental data are presented by black dots; interpolation curve is drawn by red line. Optimal threshold (0.3) is marked by red dot. **C, E**. ROC curve and its table presentation similar to panels B and D, but measurements that are highlighted by red and green rectangles on panel A were re-evaluated by two independent observers and Redness score were corrected by majority voting (see Suppl. Figure 1). Sensitivity and specificity at optimal threshold are highlighted on panels **D** and **E** by cyan color.

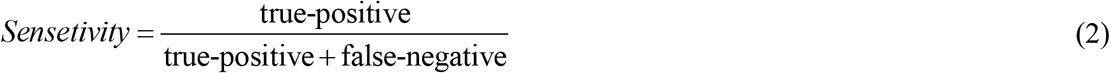

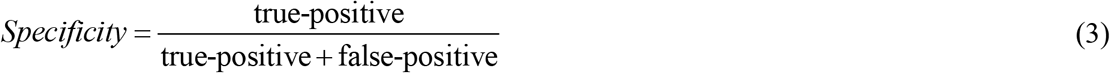

The result is drawn as black dots on Figure 3b and presented in the table on Figure 3d. The ROC-curve was smoothed (red line on Figure 3b) and the optimal threshold value was found to be 0.3 as denoted by red dot on graph Figure 3b, highlighted on table Figure 3d and is shown as a horizontal line on Figure 3a.

We found that 15 AllergoEye measurements were labelled as “false” at optimal threshold (red and green bars on Figure 3a, Suppl. Fig.1). The human-estimated scores of these measurements were re-evaluated by three independent experts. Re-evaluation changed human-estimated scores in 9 cases (marked green in Suppl.Fig.1) while 6 were confirmed (marked red in Suppl.Fig.1). In summary, AllergoEye was found to be more accurate than the human operator (6 mistakes vs. 9 mistakes respectively). After re-evaluation a new ROC-curve was build (Figure 3c,e). From table on Figure 3e, we found that AllergoEye sensitivity towards eye redness is as high as 97.7% with 90% specificity.

Since usually an allergic reaction is estimated as the sum of subjective patient’s symptoms score and objective redness score [18] (SSS), we decided to compare the AllergoEye score with SSS as a ground-truth. The scatter-plot of 114 measurements with SSS scores (from 0 to 12) on x-axis and AllergoEye score (from 0 to 3.7) on y-axis is drawn on Figure 4a by blue dots. Similar to Figure 3a-c, the ROC-curve was plotted on Figure 4b, c. Interestingly, the optimal threshold value was found to be the same (0.3) as in the redness only based ground-truth. However, the sensitivity and specificity were lower (∼87%) due to the presence of non-redness related subjective symptoms scores.

**Figure 4.**
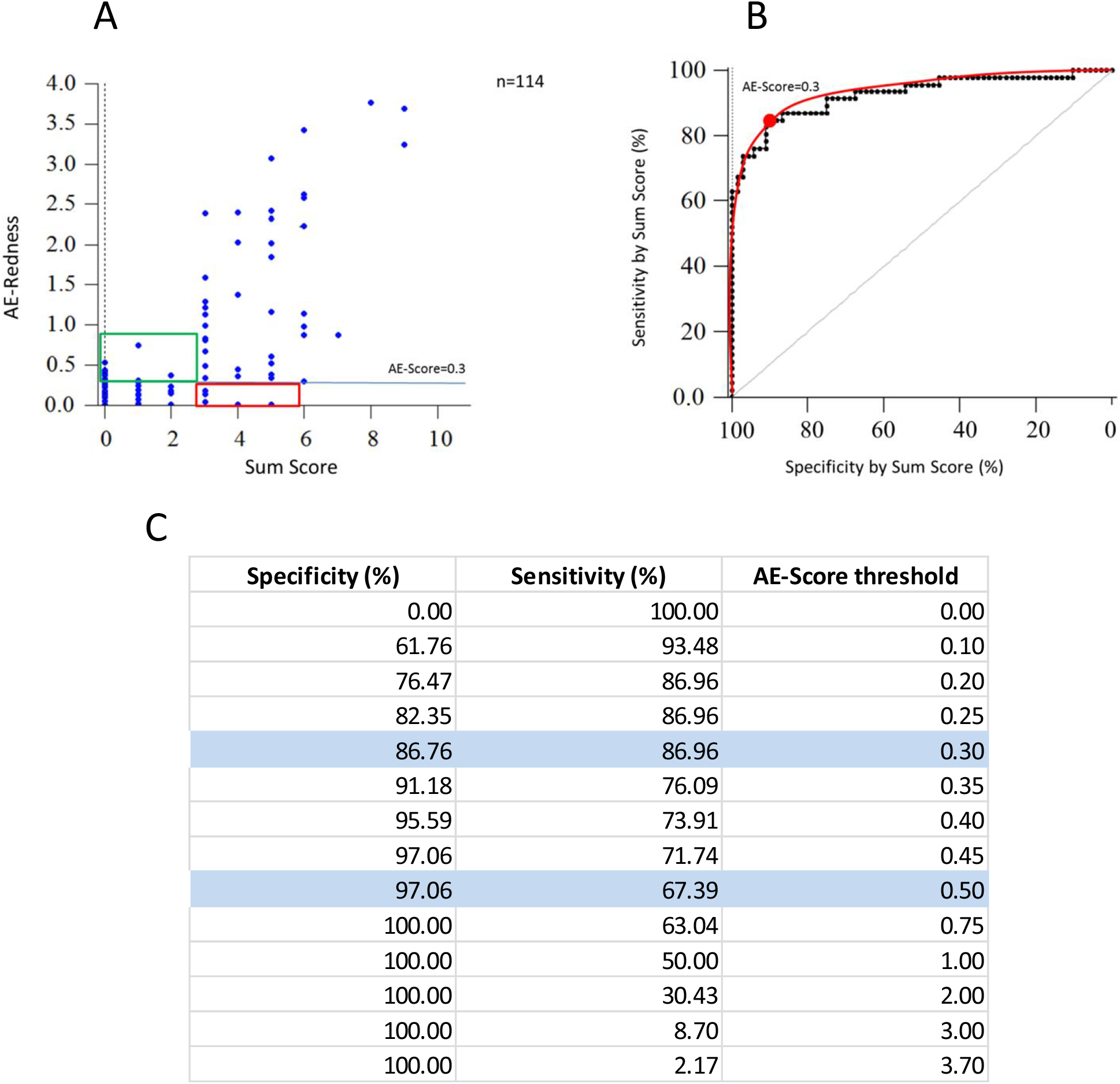
**A**. Scatter plot of 114 measurements (blue dots), which were performed on 41 patients to reveal dependency between sum symptoms score (horizontal axis) and redness evaluated by AllergoEye (vertical axis). Horizontal cyan line denotes threshold equal 0.3. Red and green rectangles highlight measurements that “false negative” and “false positive” respectively. **B, C**. ROC curve (**B**) (Sensitivity vs. Specificity) as defined by threshold value gradually changing from 0.0 to 3.7 and its table presentation (**C**). Experimental data are presented by black dots; interpolation curve is drawn by red line. Optimal threshold (0.3) is marked by red dot. Sensitivity and specificity at optimal and stringent thresholds are highlighted on panels **C** by cyan color.

In order to find the objectiveness of AllergoEye, we decided to compare the patient’s sensitivity as determined by the AllergoEye score with immunoglobulin sIgE concentration in their blood. The sIgE concentration was scored in CAP-classes [23]. The characterization of patients in the study by CAP-classes is shown on Figure 5a. Most of patients have CAP-class 3 and 4, however, 2 patients have CAP-class 6 and 2 patients have CAP-class 0 (sIgE is slightly below the low boundary of CAP-class 1). We divided patients by sensitivity category according to dilutions where the allergic reaction was detected according to criteria SSS 2≥ 3 or human-evaluated redness score > 1. For each category, the mean CAP-classes were calculated. In line with previous reports [24, 25], we did not find significant correlation between mean CAP-classes and patient’s allergic sensitivity as measured by SSS (Figure 5b). Then we built graphs for mean CAP-classes vs. AllergoEye-evaluated allergy sensitivity for the threshold value 0.3 (Figure 5c) and threshold value 0.5 (Figure 5d). Surprisingly, even with AllergoEye threshold value 0.3 we found statistically significant difference between CAP-classes of sensitive and insensitive patients (Figure 5c marked by stars). With AllergoEye threshold value 0.5 (that corresponds to a specificity 97% to SSS, Figure 4c) we revealed a statistically significant dependency between CAP-classes and patient’s allergy sensitivity (Figure 5d, marked by stars).

**Figure 5.**
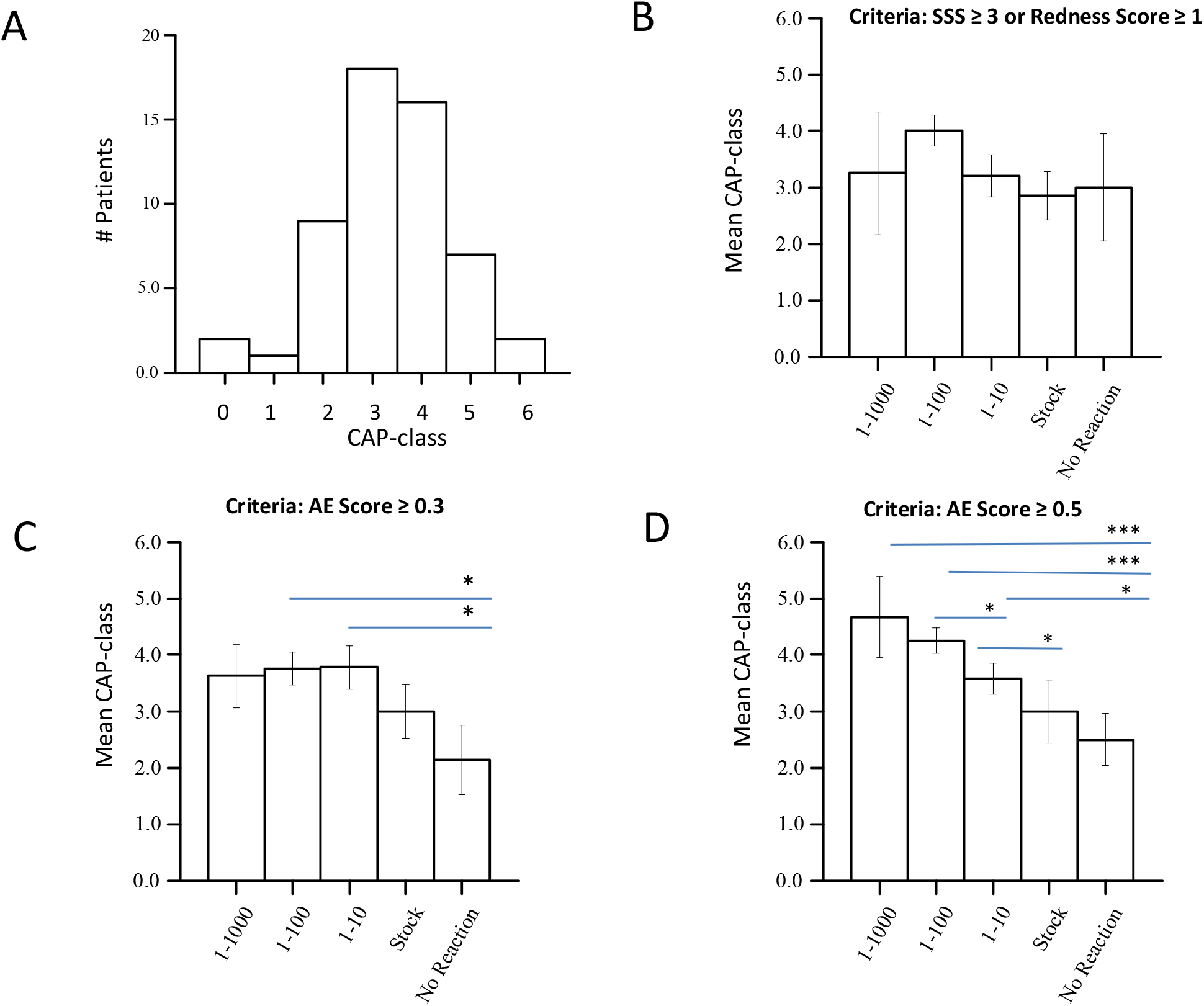
**A**. Distribution of 41 patients by sIgE CAP classes. Two patients although have CAP-class equal zero (sIgE was slightly below low boundary of CAP-class 1), but have clinically relevant symptoms. **B**. Mean CAP-class vs. sensitivity to allergen dilutions, as determined by SSS. **C**. Mean CAP-class vs. sensitivity to allergen dilutions, as determined by AllergoEye with cut-off threshold equal 0.3. **D**. Mean CAP-class vs. sensitivity to allergen dilutions, as determined by AllergoEye with cut-off threshold equal 0.5. Asterisks denote statistically significant differences, where * < 0.05; ** < 0.01; *** < 0.001.

## 4. Discussion

Recent breakthrough in deep learning resulted in wide application of artificial intelligence (AI) methods in engineering, science and medicine. Most successful AI applications were achieved in the field of image analysis (e.g. radiology, diagnostics etc.). However, AI application in allergology is rather limited.

The quantitative analysis of CPT in clinical practices is a long standing challenge in allergology. We addressed this challenge by AI-based methods for image analysis and a new method of eye redness evaluation. Up to date, most attempts to quantify the results of the CPT were based on the quantitative analysis of eye reactions on allergen in the manually segmented region of interest (ROI) in the sclera, either by measuring the apparent area of vessels [4, 12] or by per-pixel redness evaluation [4, 14]. Although succeeding in clinical studies, the methods based on manual ROI selection was too labor intensive for wide practices and methods based on the apparent area of vessels was insensitive to the diffused redness reaction of the sclera. Up to date, by best of our knowledge, only the work of Sirazitdinova with co-authors was published where automated sclera segmentation was implemented (by random forest method) [14]. However, similarly to previous works [4, 18, 12, 15] the method of Sirazitdinova et al. [14] requires matching automatically selected key points on sclera before and after provocation, which makes it too complicated for general practice usage. Indeed, such matching requires an accurate repetitive eye ball orientation from the patient during image acquisition, which significantly complicates the process of measurement and in a case of operator carelessness could lead to a significant degradation of CPT quantification accuracy.

Further, the measuring of the sclera redness is also impeded by the variation in illumination and white balance in digital cameras. During the allergen titration time that can take tens of minutes, the environment illumination conditions could change. Additional devices like a facemask were very useful, but unfortunately do not allow to totally suppress the white balance fluctuations. None of the previously proposed methods handle the changes in environment illumination conditions in an automated manner [4, 12, 14, 15].

We developed a method that uses 1) a deep neuronal network (33 layers) for automated segmentation of iris and sclera on the image, which exclude manual ROI selection and as such increases the throughput of the method; 2) a new formula (1) for brightness-independent redness per-pixel evaluation; 3) sclera redness score based on pixels redness distribution which does not require keypoints matching before and after provocation; 4) an automated white balance correction based on the untreated eye as an inner in-image control that makes the method robust towards changes in environment illumination conditions. It is worth repeating, that our method of allergic reaction evaluation is not based on widening macroscopically recognizable vessels only, but also captures changes in a capillary network that manifest itself in the diffuse redness of sclera.

The image processing and redness evaluation takes ∼15 sec. on the PC (i7, Windows 10). In rare cases, when the AI fails in an accurate segmentation of sclera, manual correction of ROI with recalculation can be performed in a few seconds. The software stores subjective symptoms and human-based redness evaluation of redness in the relational database. Cases with contradiction between human-based redness evaluation and AllergoeEye scores are easy to find by database query and re-evaluate if it is necessary.

Nowadays, the gold standard of rating the results of CPTs is the so-called summation symptom score (SSS), although there are more than one of score definitions that are used today [18]. However, the SSS includes subjective feelings of patients and fails to reveal a correlation with titer of sIgE as it was previously reported in literature [24, 25], where correlation was found for very high sIgE titers and high SSS only. Such an absence of correlation between CAP-classes and SSS was confirmed in our study (Figure 5b). One of possible explanations of absence of the correlation between SSS and CAP-classes for middle sensitive patients may be the personal ability to tolerate unpleasant feelings which differ between patients that result in wide variation in subjective score values. Therefore, SSS based methods only partially satisfy the requirement for evidence-based medicine, where especially accurate numerical values are essential in clinical studies. In contrast, the AllergoEye scores revealed an evident correlation between sIgE CAP-classes and patients’ sensitivity to allergen provocation (Figure 5d). Therefore, AllergoEye scores alone, as well as incorporated into SSS [18], could provide an objective method for clinical studies and control therapy in general allergological practice.

Validating diagnostic methods in medicine is an essential task in new developments. The main objective of this study was the validation of the AllergoEye system by determining the sensitivity and specificity curve, the so-called ROC curve, and the selection of optimal and strict thresholds for allergic response evaluation. This validation was based on two considerations. First, we compared the dilution of the CPT solution that triggered a reaction as detected by the redness determined by medical staff vs. redness detected by AllergoEye (Figure 3b,c). Second, we compared the dilution of the CPT solution that triggered reaction as detected by the redness determined by SSS vs. redness detected by AllergoEye (Figure 4b). Both comparisons showed a high sensitivity and specificity of AllergoEye (Figure 4b-d; Figure 5 b,c) as an instrument for redness and allergy assessment. In addition, the superiority of the AllergoEye over human and SSS estimation in terms of accuracy of obtained results in terms of false positive and negative results was clearly demonstrated.

Selecting the right patients for the clinical trials in allergology overall and also using CPT is an outstanding problem. The challenge is the large variability of the subjective symptoms of CPT and absence of a clear correlation between these symptoms and objective parameters, in particular the sIgE titer. Based on the AllergoEye ROC curves, the required specificity can be chosen from tables presented in Figures 4 and 5. The strict specificity allows to objectively select high sensitivity patients. At the same time the sensitivity value allows to estimate the number of patients that have to be screened which is essential for clinical study design. In a clinical trial AllergoEye could be used for quantitative monitoring of the efficiency of allergy therapy, e.g. hyposensitization.

Thus, we presented a first fully automated AI based system for quantitative evaluation of CPT results. Besides of allergology this platform could potentially be also applied for other medical areas, e.g. ophthalmology, which is also accompanied by redness of the sclera. Although this clinical validation of the method on patients with grass allergy was successful, further studies are required on a larger cohort of patients for different types of seasonal and perinatal allergies to demonstrate the universality of the AllergoEye method and its usefulness for a wider range of routine examinations and clinical studies.

## 5. Conclusion

AllergoEye, an artificial intelligence-based platform for the quantification of conjunctival provocation tests, showed high sensitivity and specificity compared to both the RMS, the redness of eye as measured by medical personnel (> 95%) and the SSS score, which represents subjective patient reports (> 86 %). It showed a clear and statistically significant correlation of the invoked allergic reaction in the eye with the IgE-CAP classes in the blood. Clinical validation demonstrated that AllergoEye is a sensitive and efficient instrument for objective evaluation of allergic reactions. It could be used for patient selection and controlling the treatment efficiency in clinical studies as well as for diagnostic and therapy control in routine allergological practice.

## Data Availability

All data produced in the present study are available upon reasonable request to the authors

## 6. Acknowledgments

We thank Dr. Andreas Bilstein for useful discussions and help in preparing clinical study, Laboratorios LETI S.L. Madrid for CPT reagents, and Alexander Kalaidzidis for software development.

The authors declare no competing financial interests.

## Author Contributions

Conceptualization Y.Yarin, Y.Kalaidzidis and Ralf Mösges; Clinical study Y.Yarin and K. Bodrova; Documentation A.Kalaidzidi; Statistical analysis Y.Kalaidzidis.

## Legends

**Suppl. Figure 1.**
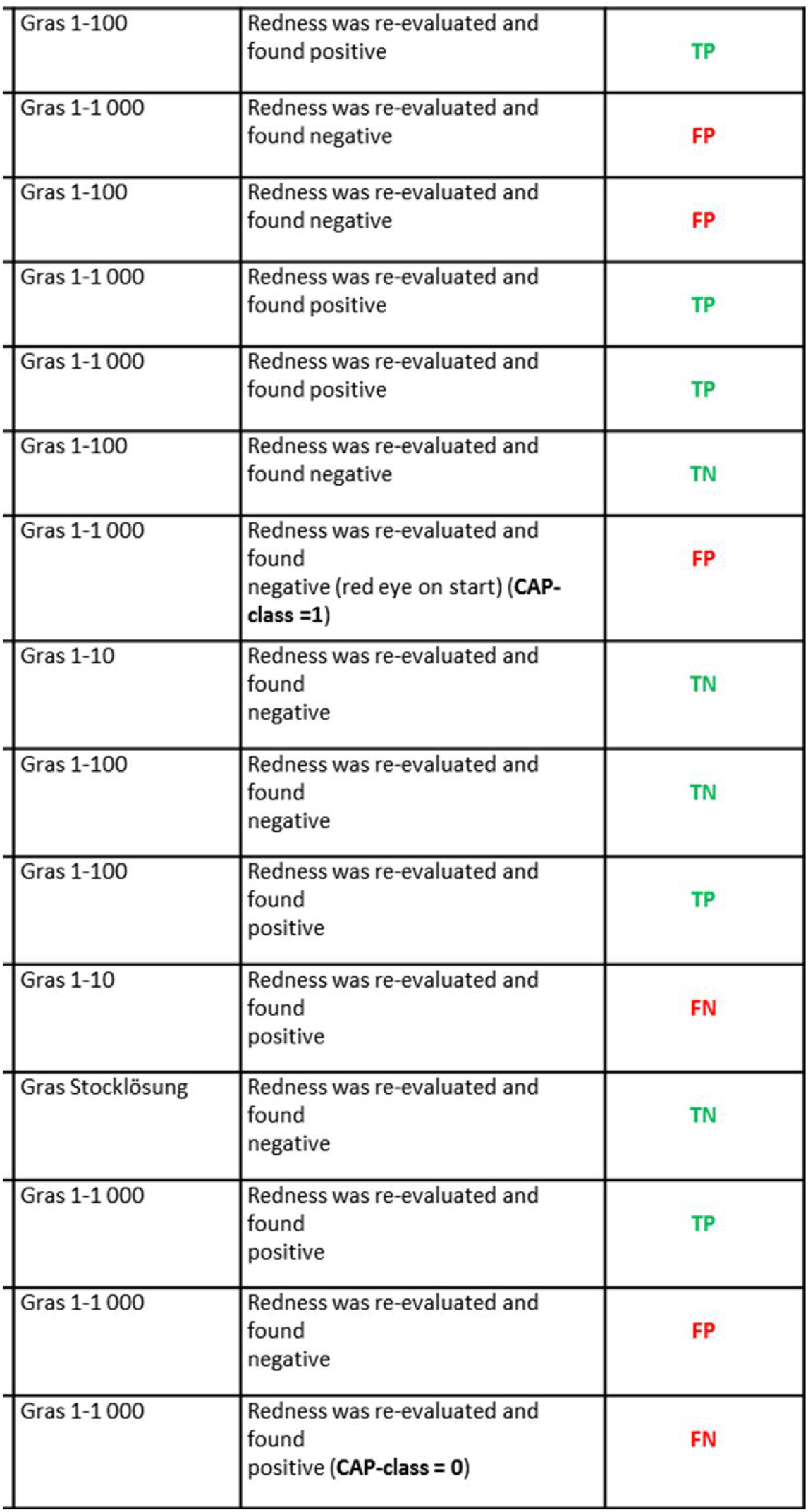
Table of re-evaluation of eye redness as defined by medial staff during CPT for the measurements highlighted on Figure 3A. The redness scores that were kept unchanged is highlighted by red, those that were changed after re-evaluation is highlighted by green. The re-evaluation was performed by 2 independent observers and final decision was done by majority votes.

**Suppl. Figure 2.**
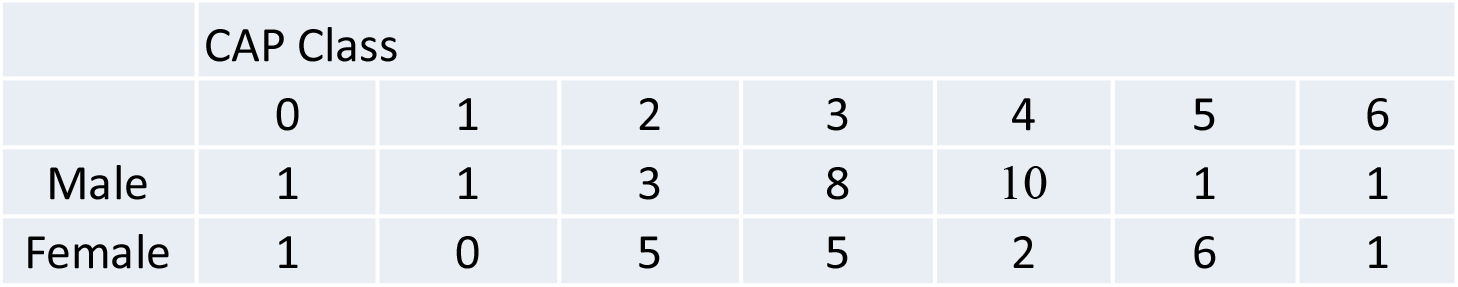
CAP-classes distribution among patients.

**Suppl. Figure 3.**
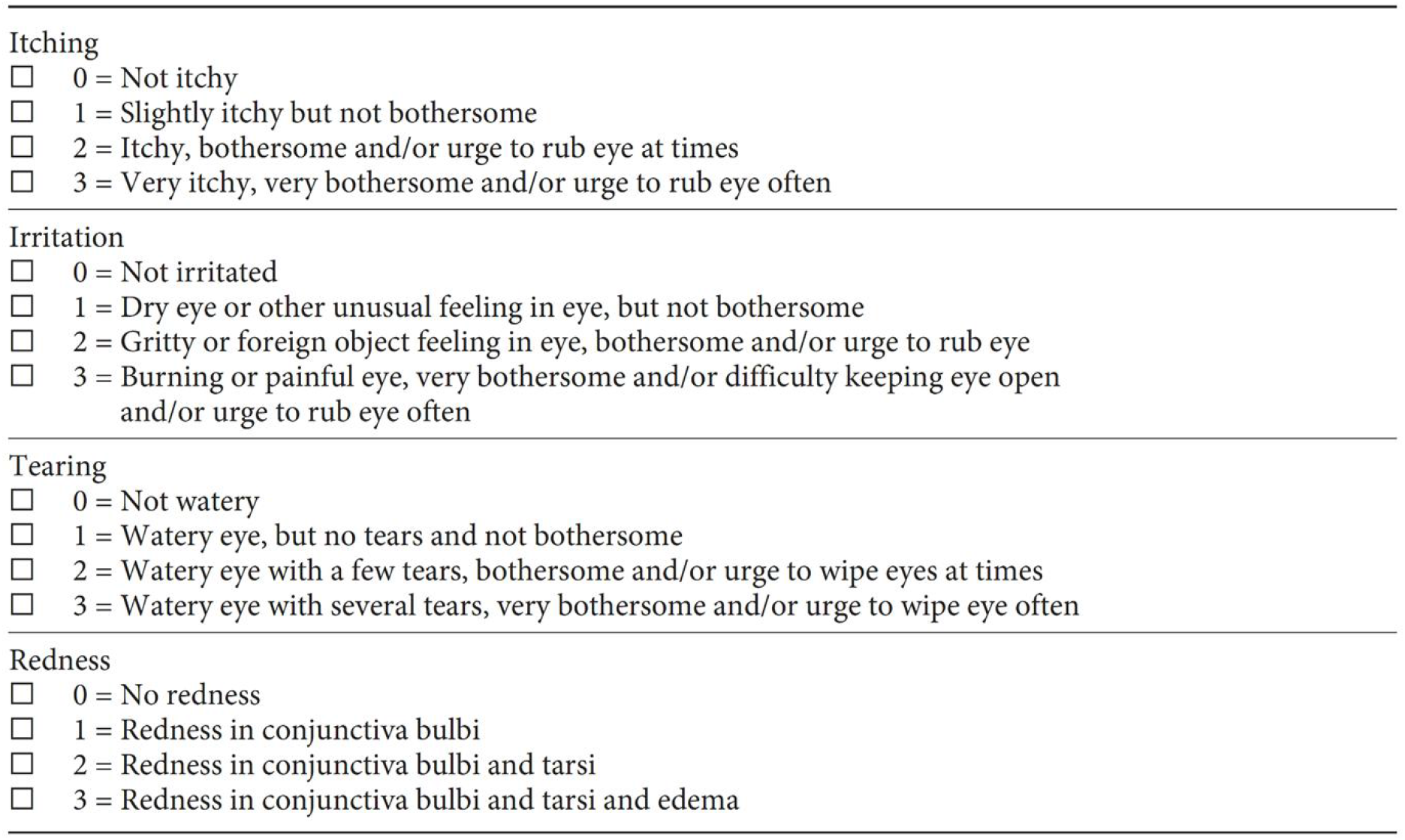
Example of symptom score evaluation protocol.

